# Mendelian randomization revealing the protective effect of sodium-glucose cotransporter 2 inhibition on prostate cancer with verified evidence from electronic healthcare and biological data

**DOI:** 10.1101/2023.10.10.23296790

**Authors:** Jie Zheng, Jieli Lu, Jiying Qi, Qian Yang, Huiling Zhao, Haoyu Liu, Youqiong Ye, Min Xu, Yu Xu, Tiange Wang, Mian Li, Zhiyun Zhao, Ruizhi Zheng, Shuangyuan Wang, Hong Lin, Chunyan Hu, Celine Sze Ling Chui, Shiu Lun Au Yeung, Shan Luo, Olympia Dimopoulou, Padraig Dixon, Sean Harrison, Yi Liu, Jamie Robinson, James Yarmolinsky, Philip Haycock, Jinqiu Yuan, Sarah Lewis, Tom R. Gaunt, George Davey Smith, Ning Guang, Richard M. Martin, Bin Cui, Weiqing Wang, Yufang Bi

**Author notes:** Equal first author (Jie Zheng, Jieli Lu). **Senior and corresponding author** Yufang Bi, Professor of medicine, Department of Endocrine and Metabolic Diseases, Shanghai Institute of Endocrine and Metabolic Diseases, Ruijin Hospital, Shanghai Jiao Tong University School of Medicine, Shanghai, China, Weiqing Wang, Professor of medicine, Department of Endocrine and Metabolic Diseases, Shanghai Institute of Endocrine and Metabolic Diseases, Ruijin Hospital, Shanghai Jiao Tong University School of Medicine, Shanghai, China, Richard M. Martin, Professor of Clinical Epidemiology, MRC Integrative Epidemiology Unit (IEU), Bristol Medical School, University of Bristol. Population Health Sciences, Bristol Medical School, University of Bristol. NIHR Biomedical Research Centre at the University Hospitals Bristol and Weston NHS Foundation Trust and the University of Bristol, United Kingdom., Jie Zheng, Professor in health informatics, Department of Endocrine and Metabolic Diseases, Shanghai Institute of Endocrine and Metabolic Diseases, Ruijin Hospital, Shanghai Jiao Tong University School of Medicine, Shanghai, China.

## Abstract

**Background:** Observational studies indicated a decreased risk of prostate cancer by SGLT2 inhibitors, but high-quality evidence is lacking to make a clear conclusion. We evaluated the effect of SGLT2 inhibition on prostate cancer risk by triangulating evidence from three methods.

**Methods:** Genetic variants associated with HbA_1c_ levels (P<5×10^-8^) in the genomic region of the target gene, *SLC5A2*, were used to proxy SGLT2 inhibition. In discovery, Mendelian randomization (MR) was applied to estimate effects of genetically proxied SGLT2 inhibition on risk of prostate cancer and its subtypes (79,148 cases and 61,106 controls). In a validation using electronic healthcare data, the association of incidence of prostate cancer between 24,155 new users of SGLT2 inhibitors and 24,155 new users of the active comparator, dipeptidyl peptidase 4 inhibitors, was estimated using electronic health-care data. In a biological validation, the differential gene expression of *SLC5A2* between normal prostate tissue and tumour tissue were estimated in 691 prostate cancer patients. To validate the influence of glucose, the association between HbA_1c_ levels and incident prostate cancer during 10-years of follow-up were estimated.

**Findings:** For genetic evidence, genetically proxied SGLT2 inhibition reduced the risk of overall (odds ratio=0.56, 95%CI=0.38 to 0.82), advanced (OR=0.52, 95%CI=0.27 to 0.99) and early-onset (OR=0.27, 95%CI=0.11 to 0.72) prostate cancer. For electronic healthcare evidence, usage of SGLT2 inhibitor was associated with a 23% reduced risk of prostate cancer (hazard ratio=0.77, 95%CI=0.61 to 0.99) in males with diabetes. For biological evidence, expression levels of the *SLC5A2* gene in tumour prostate tissue was 2.02-fold higher than that in normal tissue (P=0.006). Genetically proxied HbA_1c_ and observed HbA_1c_ provided little evidence to support an association with total/incident prostate cancer, implies a non-glycemic effect of SGLT2 inhibition on prostate cancer.

**Interpretation:** This study provides genetic, electronic healthcare, and biological evidence to support a beneficial effect of SGLT2 inhibition on reducing prostate cancer risk. Future trials are warranted to investigate whether SLGT2 inhibitors can been recommended for diabetic individuals with high risk of prostate cancer or considered as an anti-prostate cancer therapy.

**Funding:** AMS, MRC and NSFC.

**Graphical abstract:** 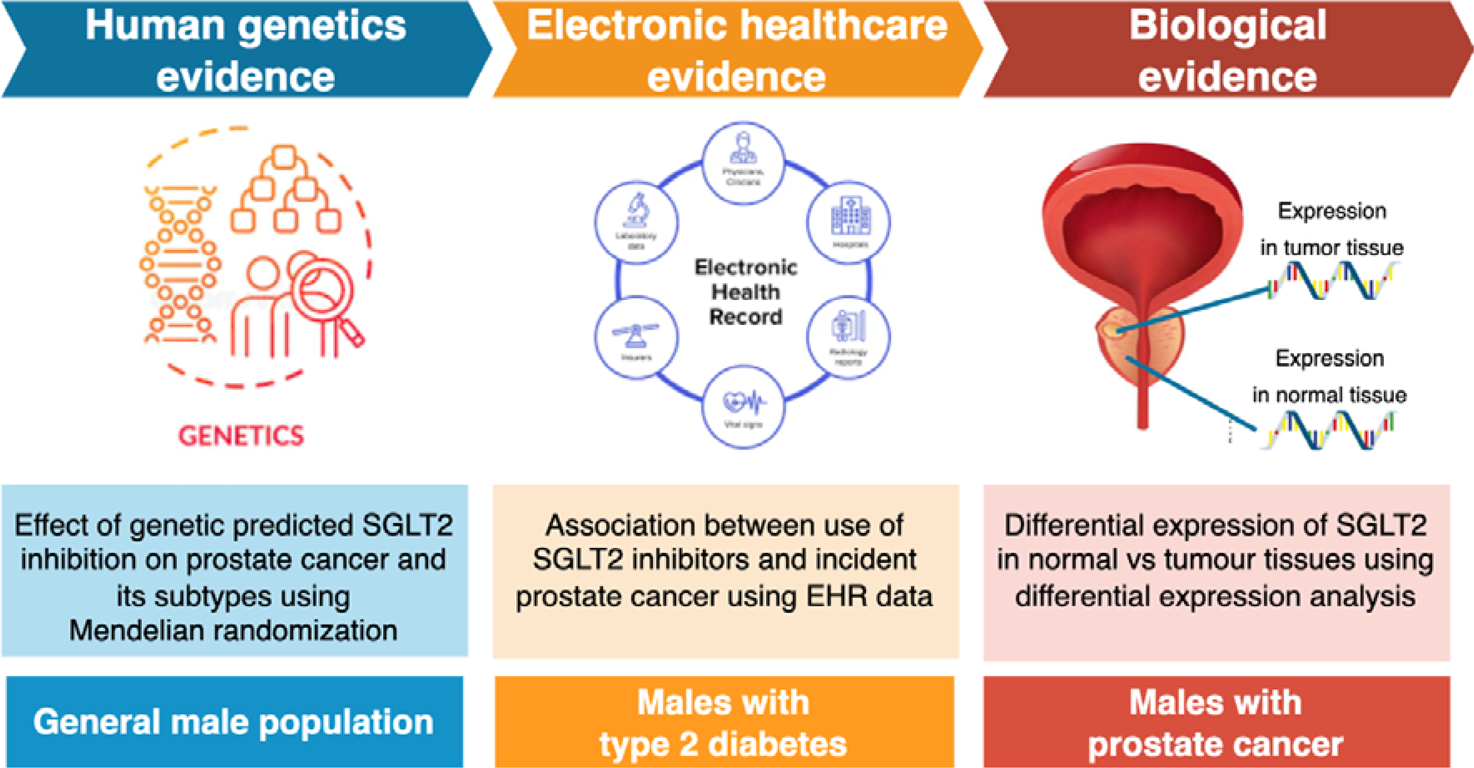

**Research in context:** *Evidence before this study:* We searched PubMed, Embase and clinicaltrials.gov databases from inception up to July 11, 2023 using the search terms: “SGLT2 inhibitor”, “canagliflozin”, “dapagliflozin”, or “empagliflozin” and “prostate cancer” and “clinical trials”, without language restrictions. Some functionals studies provided evidence of SGLT2 inhibition on reduce the viability of prostate cancer cells but lack of human-based evidence. Only one clinical trial study is investigating the role of SGLT2 inhibitors on prostate cancer in individuals with diabetes. Other 46 trials of SGLT2 inhibitors set prostate cancer as secondary outcome, the prostate cancer cases were limited for these studies, power issues have prevented clear causal inference. Little has been done to establish the causal role of SGLT2 inhibition on total and incident prostate cancer.

*Added value of this study:* In this study, Mendelian randomization (MR) was applied in 140,254 men (79,148 with prostate cancer), and suggested that genetically proxied SGLT2 inhibition showed an effect on 44%, 48% and 73% reduced risk of total-, advanced- and early-onset prostate cancer in the general male population. Validation analysis using electronic health-care record data (81,122 men with diabetes) suggested that usage of SGLT2 inhibitor was associated with a 23% reduced risk of prostate cancer in males with diabetes. The differential expression analysis in 639 men with prostate cancer showed that the expression of SGLT2 was 2.02 folds higher in prostate cancer tissues compared with that in surrounding normal prostate tissues. As a benchmark, MR and observational analyses showed little evidence to support an effect of HbA1c on prostate cancer, which suggests a potential non-HbA_1c_ effect of SGLT2 inhibition on prostate cancer.

*Implication of all the available evidence:* There were multiple sources of evidence to support a protective role of genetically proxied SGLT2 inhibition and usage of SGLT2 inhibitors on risk of prostate cancer in men with and without diabetes and/or prostate cancer. Future clinical trials should be prioritised for investigation of the long-term use of SGLT2 inhibitors in the prevention and treatment of prostate cancer.

## Introduction

Diabetes is one of the most common chronic conditions that affecting 537 million individuals in 2021^1^. Among various types of anti-diabetic drugs, recent clinical trials showed the beneficial effect of sodium-glucose cotransporter 2 (SGLT2) inhibitors on reduced risk of atherosclerotic cardiovascular disease (ASCVD) in addition to improvements in HbA_1c_^2–4^. Based on the trial evidence, the American Diabetes Association (ADA) and European Association for the Study of Diabetes (EASD) guidelines recommended SGLT2 inhibitors as first-line therapy for patients with or with indicators of high risk of ASCVD, heart failure, or chronic kidney disease (CKD) since 2020^5,6,7^. It has now been widely used by clinicians from endocrinology and cardiology departments.

Cancer was known to be one of the common comorbidities for type 2 diabetes^8,9,10^. Among various cancer types, prostate cancer is the second most commonly diagnosed male cancer, with nearly 1.41 million new diagnosis worldwide in 2020, and is a major cause of cancer death in men^11^. However, no clinical guideline recommended usage of anti-diabetic drugs on individuals with cancers or in high risk of cancers, especially for males with these two common diseases, diabetes and prostate cancer. A recent review summarized the anti-cancer mechanisms of SGLT2 inhibitors^12^. Observational studies have also reported a decreased risk of prostate cancer amongst men with diabetes who are taking SGLT2 inhibitors^13^. However, the largest meta-analysis of randomized controlled trials (RCTs) in individuals with T2DM suggested little difference in prostate cancer incidence between users of SGLT2 inhibitors and users of placebo or active comparators^14^, although in this study power could be an issue due to limited number of incidence prostate cancer cases (N=41) included in the analysis. Correctively, existing epidemiology studies provided some clues but the evidence level to support the protective effect of SGLT2 inhibition on prostate cancer risk was still low. Whether SGLT2 inhibition can be recommended for diabetic individuals with high risk of cancers or can be even repurposed as an anti-cancer drug target need further investigation.

Evidence triangulation is the practice of obtaining more reliable answers to research questions through integrating results from several different methods^15^. These methods have different assumptions and unrelated sources of biases. If results of these methods point to a similar conclusion, this will strengthen confidence in the finding. For the causal question of identifying the effect of a drug target on a disease, human genetics, epidemiology and bioinformatics are three commonly used approaches^16,17,18,19,20,21^. Triangulating evidence from these methods in one study may provide an attractive strategy to improve evidence level for drug repurposing. Mendelian randomization (MR) is a method that uses germline genetic variants as proxy measures of exposure and estimates the causal effect of an exposure on an outcome^22^. An individual’s germline genetic make-up influences their biology from conception, meaning that causal estimates from MR studies reflect lifelong exposures (e.g. lifelong SGLT2 inhibition) and that the estimates are not generally susceptible to reverse causation or confounding^23^. Observational associations of the use of a drug on disease incidence are normally estimated using Cox proportional hazard models, where a ‘new user active comparators’ design may reduce the influence of confounders^24^. Differential expression analysis is a commonly used approach to estimate the association of expression of a target gene on a disease, which is commonly used to identify genetic mechanisms for cancers. Due to enriched data resources to support application of all three methods^25,26,27^, studying the effect of SGLT2 inhibition on prostate cancer is a preferred example for such evidence triangulation strategy.

The objective of this study was to estimate the causal effects of SGLT2 inhibition on prostate cancer and its subtypes by triangulating evidence from human genetics, electronic healthcare and biological data. The effect of HbA_1c_ on prostate cancer were further estimated using human genetics and observational epidemiology approaches.

## Methods

### Summary of study design and data sources

The **graphical abstract** and **Figure 1** present an overview of three sets of analyses conducted in this study. Each analysis aims to answer the same causal question in different subpopulations. All studies contributing data to this analysis had the relevant institutional review board approval from each country and all participants provided informed consent.

**Figure 1.**
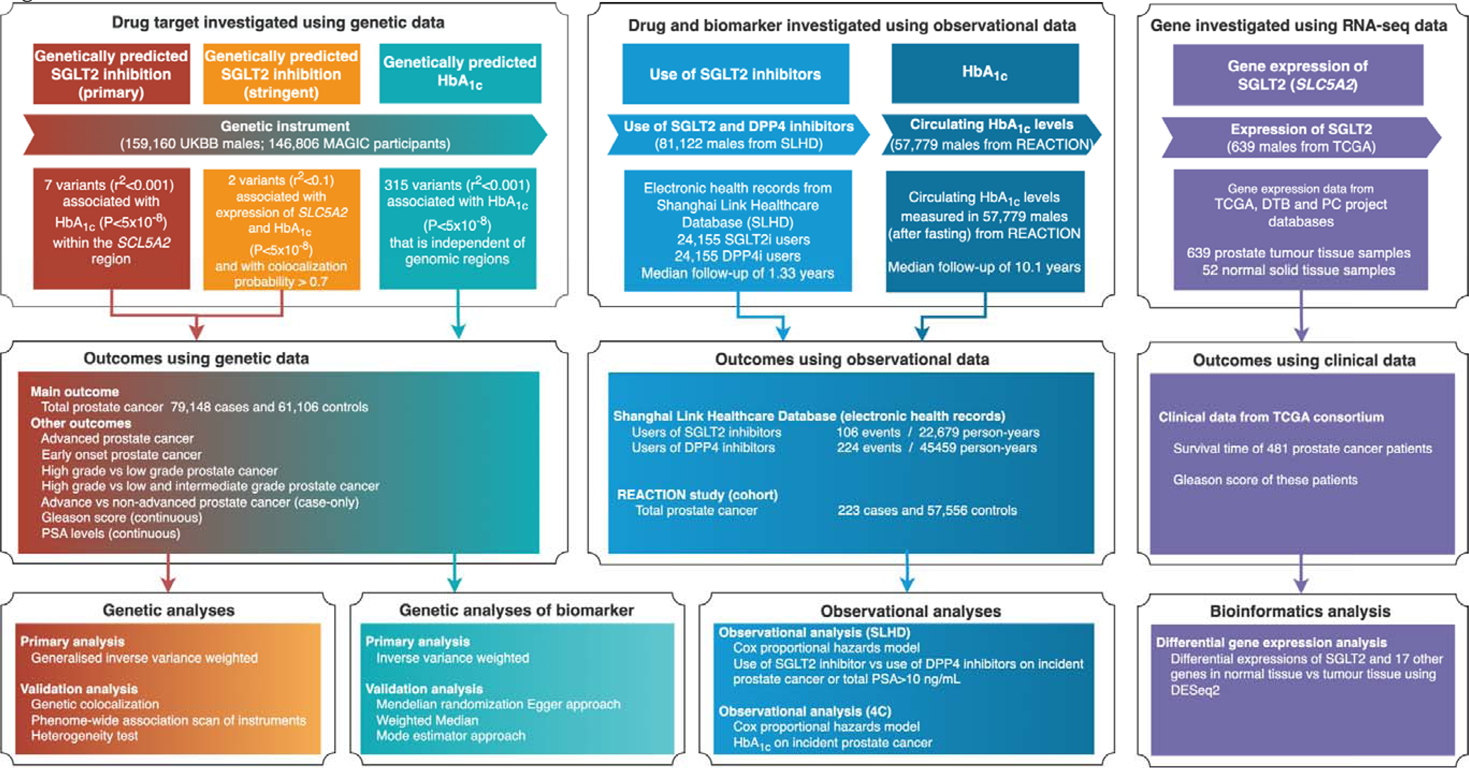
Genetic instrument selection, data sources, and analysis strategy in a triangulation study of the effect of SGLT2 inhibition on prostate cancer. For human genetic analyses, the effect of sodium–glucose cotransporter 2 (SGLT2) inhibition on risk of prostate cancer and its subtypes were estimated using Mendelian randomization. For observational analyses, the effect of use of SGLT2 inhibitors on incident prostate cancer risk were estimated in males with diabetes. Use of DPP4 inhibitors were used as an active comparator. For computation biology analyses, the differential expression of SGLT2 in prostate tumour tissue vs normal prostate tissue was estimated. Then the association of SGLT2 expression on prostate cancer progression (proxied by Gleason score) and survival time were estimated. More details of instrument selection and analysis strategies were listed in **Appendix Note 1-4**.

First, the human genetics analysis was applied as the discovery analysis in the general male population. We estimated the putative causal effects of SGLT2 inhibition and genetically predicted HbA_1c_ on risks of prostate cancer and its subtypes using MR. The summary genetic association data from a case-control genome-wide association study (GWAS) of prostate cancer in the PRACTICAL and GAME-ON/ELLIPSE Consortium^25,26^ were used (N=140,254 men from the general population).

Second, as a validation of the MR findings, two sets of analyses were conducted using two independent large-scale cohort resources: (1) the association of use of SGLT2 inhibitors on incident prostate cancer were estimated in diabetic individuals using data derived from electronic health record data in the Shanghai Link Healthcare Database (SLHD; N=81,122 men with diabetes), a representative clinical database covering electronic health-care records for over 99% of Shanghai residents since 2013^28^; (2) the association of baseline HbA_1c_ levels with incident prostate cancer during 10-years of follow-up was estimated using data from the Risk Evaluation of Cancers in Chinese Diabetic Individuals (REACTION) study^8^ (N=57,779 men for the general population). Both human genetics and observational analyses were related to prostate cancer risk, which are related to disease prevention.

Third, the biological analysis was conducted to understand the biological function of SGLT2 expression on prostate cancer in patients with prostate cancer. The differential gene expression analysis using data from the TCGA Program^27^ was conducted (https://portal.gdc.cancer.gov/; N=639 men with prostate cancer). This analysis was conducted in prostate cancer patients, which may inform cancer treatment.

## Statistical analysis

### Discovery analyses using Mendelian randomization

#### Genetic instrument selection for SGLT2 inhibition

To generate genetic instruments to proxy the lifelong glucose-lowering effect of SGLT2 inhibition, summary data were obtained from a GWAS of HbA_1c_ levels in UK Biobank (N=344,182). Seven genetic variants robustly and independently associated with HbA_1c_ (P<5×10^-8^; square of linkage disequilibrium [LD] among instruments [r^2^] <0.1) in the *SLC5A2* region were selected as instruments to proxy SGLT2 inhibition (noted as <u>primary</u> <u>instruments</u>; **Table 1**). Since prostate cancer is male-specific, the genetic associations of the selected instruments on HbA_1c_ from 159,160 UK Biobank males were used in the MR analysis (**Appendix Table 1A**).

**Table 1.**
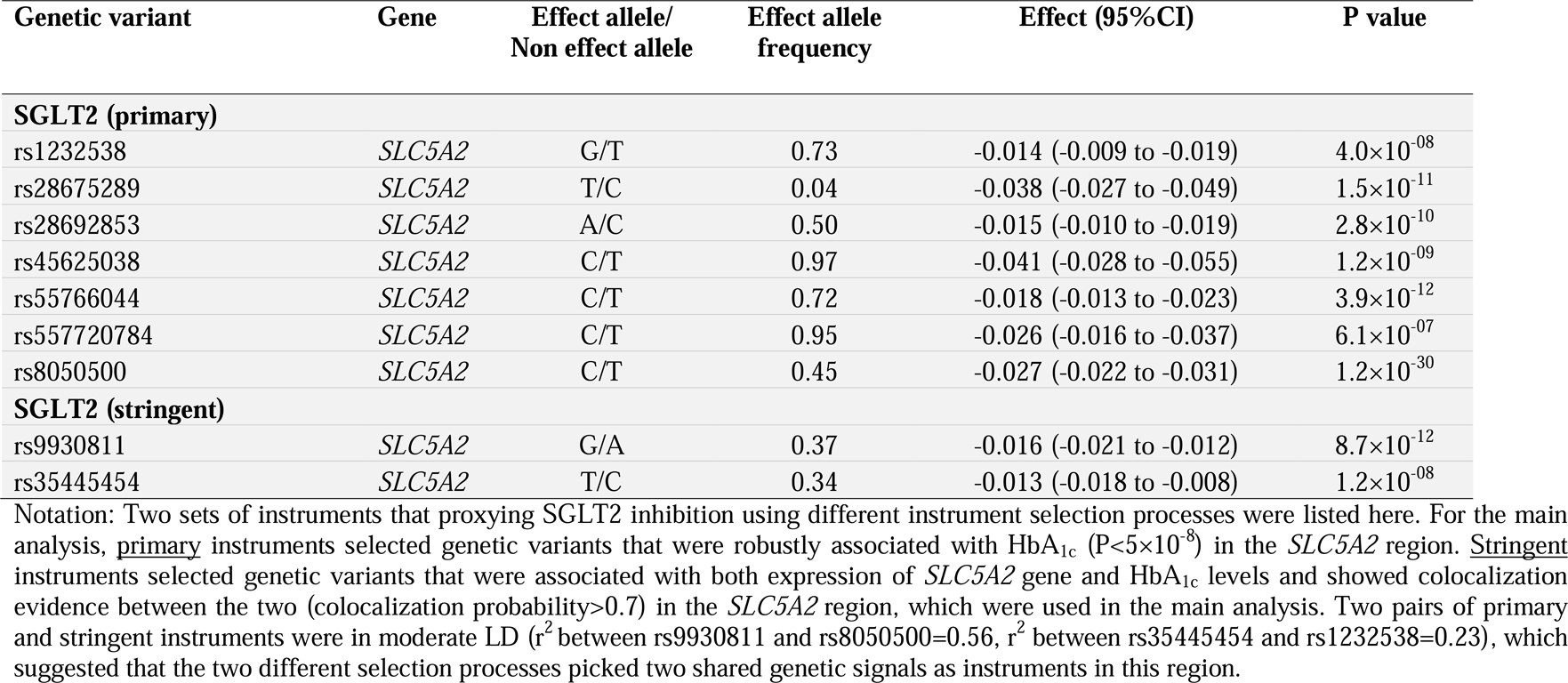
Characteristics of genetic variants associated with HbA_1c_ (per 0.62% lowering) or expression levels of *SLC5A2* gene and used as proxies for SGLT2 inhibition in the general population.

As a validation, we used a more stringent instrument selection pipeline, as described before^29,30^, which hypothesized that a genetic variant is a valid instrument for SGLT2 inhibition if this variant showed genetic colocalization evidence^31,32^ on both HbA_1c_ and expression levels of the target gene of SLGT2, *SLC5A2*^33,34^. After applying this process, two variants were selected as instruments (noted as <u>stringent instruments</u>; **Table 1**, **Appendix Table 2**).

We further validated the reliability of the instruments using an independent dataset. The genetic effects of the SGLT2 instruments were extracted from a European-only GWAS of HbA_1c_ within the MAGIC consortium^35^ (N=146,806 **Appendix Table 3**). To instrument HbA_1c_ levels, 315 independent variants (LD r^2^<0.001) associated with HbA_1c_ (P<5×10^-8^) were obtained from UK Biobank male only GWAS data (**Appendix Table 1B**).

More details of the instrument selection are described in **Appendix Note 1** and **Appendix Figure 1**).

#### Outcome selection for human genetics analysis

Eight prostate cancer related phenotypes were selected as outcomes for the MR analysis: total-, aggressive-, early-onset-, high aggressive vs low aggressive-, high aggressive vs low and intermediate aggressive-, advanced stage vs localised stage prostate cancer. Advanced prostate cancer was defined as metastatic disease or Gleason score (GS) ≥ 8 or PSA > 100 or prostate cancer death; early-onset refers to prostate cancer onset before age 55; low aggressive refers to T stage from the TNM staging ≤ T1, and GS ≤ 6, and PSA<10; intermediate aggressive refers to T stage: T2, and GS=7, and PSA 10∼20; and high aggressive refers to T stage: T3/T4 or N1 or M1 or GS ≥ 8 or PSA >20. PSA levels were included as they drive prostate cancer diagnoses, and we wanted to exclude an effect of the exposures on PSA that could bias the prostate cancer associations. Summary data were obtained from GWAS results in the PRACTICAL and GAME-ON/ELLIPSE consortia or Kachuri et al.^25,36^. Detail information of the prostate cancer related outcomes were listed in **Appendix Table 4**.

#### Mendelian randomization analyses

Germline genetic variants used to proxy SGLT2 inhibition were matched to prostate cancer datasets by orienting effects of the exposure and the outcome to the same effect allele. If an instrument was missing in the outcome dataset, a genetic variant with high LD (r^2^>0.8) to the instrument was selected as a proxy instrument where possible. An inverse-variance weighted approach was used to combine variant-level Wald ratio estimates into an overall effect estimate^37^. All MR estimates (odds ratios [ORs]) were scaled to SD unit to reflect the equivalent of a one SD unit (0.62%) reduction in HbA_1c_.

A set of Mendelian randomization In the main MR analyses, the effects of genetically proxied SGLT2 inhibition (using seven primary instruments) were estimated on total prostate cancer, its subtypes and PSA levels in the general male population (PRACTICAL and GAME-ON/ELLIPSE)^25^. The effect of SGLT2 inhibition on T2DM^38^ was estimated as a positive control analysis. For the validation MR analyses, the effects of SGLT2 inhibition on the prostate cancer related outcomes were estimated using the stringent instruments and instruments from the independent dataset (MAGIC).

We report findings according to the STROBE-MR (Strengthening the Reporting of Mendelian Randomization Studies) guidelines^39,40^ (the STROBE-MR check list attached). MR has three key assumptions: (1) the germline genetic instruments used to proxy SGLT2 inhibition are robustly associated with the exposure (“relevance”); (2) the instruments are not associated with common causes (confounders) of the instrument-outcome association (exchangeability); (3) the instruments are only associated with the outcome through the exposure under study (“exclusion restriction”). These MR assumptions were tested using the sensitivity methods, including generalized inverse variance weighted (gIVW)^41^, genetic colocalization^31,32^, phenome-wide association studies (which including classic risk factors associated with SGLT2 instruments) using data from the IEU OpenGWAS database^26^, heterogeneity tests across instruments using Cochran’s Q^42^, weighted median^43^ and mode-based estimate approaches^44^ and Multivariable MR^45^. Extended descriptions of these sensitivity analyses are provided in the **Appendix Note 3**. Moreover, the SGLT2 instruments were associated with other 17 genes. Associations between expression of these genes and prostate cancer may reflect pleitropy, but hard to be tested. Here, we estimated whether expression levels of these genes were different in normal tissue versus prostate tumour tissue using DESeq2^46^ (data from TCGA), which provided additional evidence to distinguish pleiotropy. For all MR analyses, Bonferroni corrections were applied to establish multiple testing-adjusted thresholds. All the MR analyses were conducted using the TwoSampleMR R package v0.5.6^47^.

#### Validation using electronic healthcare data and cohort with long-term follow-up

To triangulate with the MR findings, we estimated the association between the use of SGLT2 inhibitors and the incidence of prostate cancer during the follow-up period (**Figure 2A**; data from the Shanghai Link Healthcare Database [SLHD]). To emulate a trial protocol using large-scale observational data^48^, we setup stringent eligibility criteria and used a ‘new user active comparator’ design (more details in **Appendix Note 2**). To reduce the influence of potential confounders, dipeptidyl peptidase 4 (DPP4) inhibitors were selected as active comparators since it is a newer second-line anti-diabetic drug that had been compared with SGLT2 inhibitors in previous clinical trials^49^. Within SLHD, 130,817 males with T2DM newly treated with SGLT2 inhibitors (or DPP4 inhibitors) were selected. The look back period for this study was from 1^st^ of Jan 2013 till cohort entry (which was the first time a male had an outpatient visit record in any tier-three hospital in Shanghai). After selection, 81,122 males who fitted the eligibility criteria were selected as the original cohort of this study. The follow-up period during which prostate cancer diagnosis was recorded was 1^st^ of March 2017 (when SGLT2 inhibitors was marketed in China) till 31^st^ of December 2021 (median follow-up period was 1.33 years). Propensity-score matching was conducted using 30 covariates, aiming to balance the baseline characteristics between the two treatment groups and reduce confounding effects: 12 comorbidities, 7 anti-diabetic drugs and 11 other medications were used in building up the propensity score (**Appendix Table 5**). An adjusted Cox proportional hazard’s model was used and hazard ratio (HR) with 95% confidence intervals (CIs) were calculated to estimate the risk of incident prostate cancer among new users of SGLT2 inhibitors as compared with new users of dipeptidyl peptidase 4 (DPP4) inhibitors.

**Figure 2.**
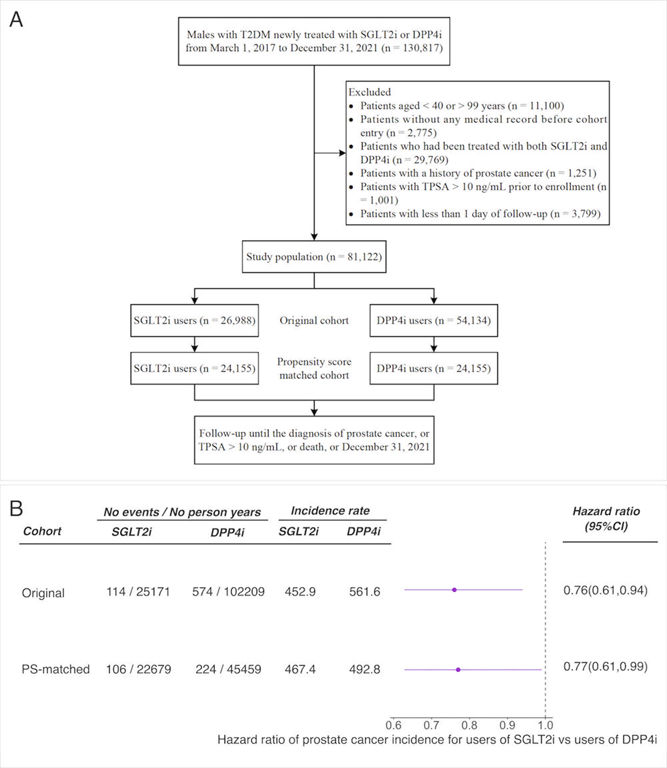
Flowchart of patient inclusion and association between use of sodium-glucose cotransporter 2 (SGLT2) inhibitors and risk of incident prostate cancer or at high risk of prostate cancer. (A) flowchart of patient inclusion in the study population. SLGT2i=sodium glucose cotransporter 2 inhibitors; DPP4i=Dipeptidyl peptidase 4 inhibitors; TPSA=total prostate specific antigen. A patient could be excluded for more than one reason. (B) The association between use of SGLT2 inhibitors compared with DPP4 inhibitors and risk of prostate cancer or with total PSA > 10 ng/mL (which indicated high risk of prostate cancer). The covariates been used in this analysis including demographic data (age); comorbidities (benign prostatic hyperplasia, hypertension, dyslipidemia, diabetic complications, ischemic heart disease, peripheral vascular disease, heart failure, cerebrovascular disease, chronic lung disease, moderate or severe kidney disease, moderate or severe liver disease, and other cancers); antidiabetic drugs (metformin, insulin, glucagon-like peptide-1 receptor agonist, sulfonylurea, glinide, α-glucosidase inhibitor, and thiazolidinedione); and other medications (angiotensin converting enzyme inhibitor, angiotensin receptor blocker, calcium channel blocker, α/β- blockers, diuretic, statin, fibrate, aspirin, other antiplatelet drugs, non-steroidal anti-inflammatory drug, and 5α-reductase inhibitor). The unit of the incidence rate was 100,000 person years.

For the second observational analysis using data from the REACTION study^8^, we defined the outcome as incident prostate cancer during the 10-years of follow-up period. Prostate cancer was defined using ICD 10 code “C61” for the two studies. We estimated the observational association between baseline HbA_1c_ and incident prostate cancer during 10 years of follow-up in the REACTION study (223 cases and 57,556 controls)^8^. HbA_1c_ levels were measured at baseline and used as a continuous variable with per SD increasement in the analysis (mean=6.04%, SD=1.11%). Age, body mass index, tobacco consumption, alcohol consumption, physical activity, and diet score were included as covariates in the Cox proportional hazards model (more details in **Appendix Note 2**).

#### Biological validation using gene expression data in cancer patients

To dig into functional mechanism, a differential gene expression analysis was carried out using DESeq2^46^ to understand the expression patterns of SGLT2 (counts) in normal prostate tissue versus prostate tumour tissue (data from GDC data portal https://portal.gdc.cancer.gov/). After selection, 691 prostate tissue samples from 639 individuals, involving 639 prostate tumour samples and 52 normal solid tissues, were selected from three databases: TCGA, West Coast Prostrate Cancer Dream Team (WCDT) and Count Me In (CMI) Project. The dataset batch was set as a covariate in the model to adjust for batch effect.

## Results

### Discovery: MR effects of SGLT2 inhibition on prostate cancer risk

The characteristics of the primary and stringent genetic instruments used to proxy SGLT2 inhibition are listed in **Table 1** and **Appendix Table 1-3,** respectively. Across these exposures, the F-statistics used to test the relevance MR assumption suggests that weak instrument bias was unlikely to be an issue in this study.

Genetically proxied SGLT2 inhibition (estimated by primary instruments), equivalent to a one SD (0.62%) reduction in HbA_1c_, reduced the risk of total prostate cancer by 44% (OR=0.56, 95%CI=0.38 to 0.82, P=0.003; **Table 2**). This effect was consistent across the seven instruments (heterogeneity P=0.80; **Figure 3**). The other four sensitivity MR models showed similar effect estimates (**Appendix Figure 2**).

**Figure 3.**
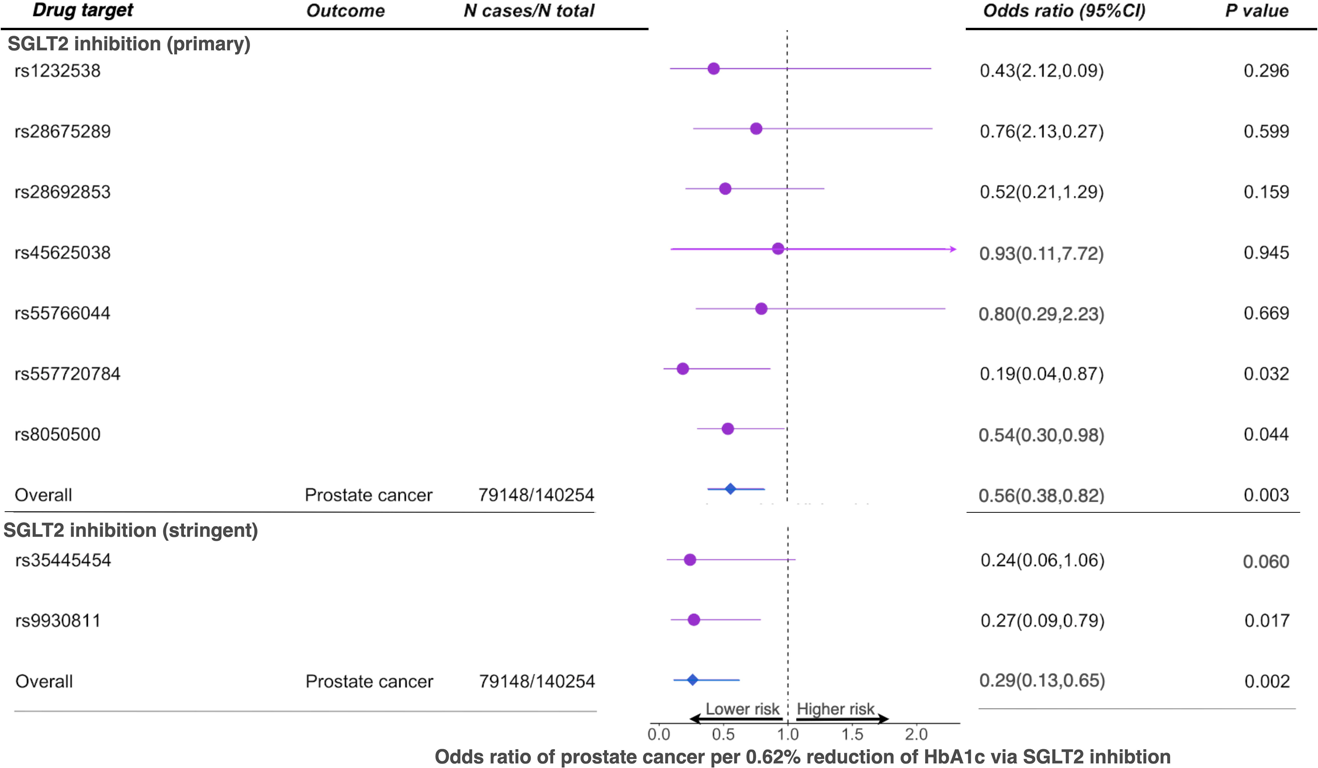
Mendelian randomization estimates of the effects of sodium–glucose cotransporter 2 (SGLT2) inhibition on prostate cancer risk in the general European population. Two sets of genetic instruments were used in this analysis. Primary instruments included seven genetic variants that were associated with HbA_1c_ (P<5×10^-8^) in the *SLC5A2* region. Stringent instruments were two genetic variants associated with both expression levels of *SLC5A2* and HbA_1c_ levels (with colocalization probability>0.7 between the two) in the *SLC5A2* region.

**Table 2.**
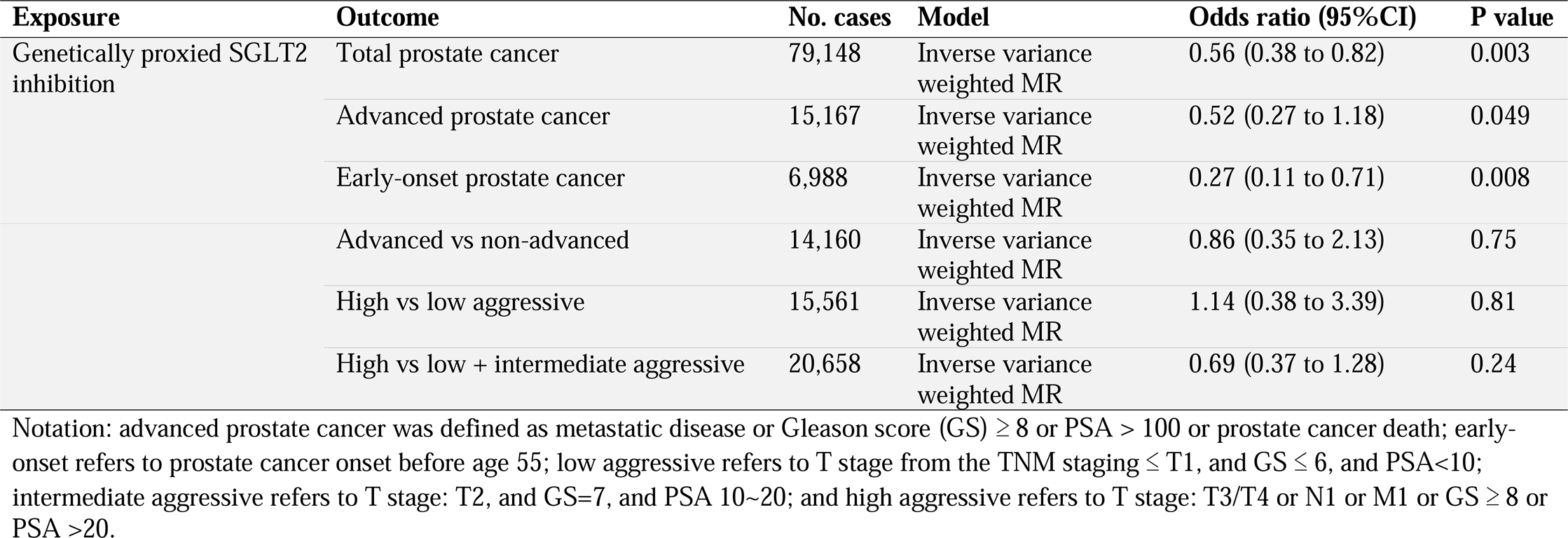
Effect estimates of genetically proxied SGLT2 inhibition and genetic proxied HbA_1c_ levels on total, aggressive and early onset prostate cancer among men in general population using data from PRACTICAL and GAME-ON/ELLIPSE consortium.

Genetically proxied SGLT2 inhibition lowered the risk of advanced (OR=0.52, 95%CI=0.27 to 0.99; P=0.049) and early-onset (OR=0.27, 95%CI=0.11 to 0.71; P=0.008) prostate cancer. Little evidence was observed to support an effect of SGLT2 inhibition on other prostate-cancer related outcomes (**Table 2**). In addition, there was little evidence to support an effect of SGLT2 inhibition on PSA levels (β=0.14, 95%CI=0.030 to −0.30, P=0.107; **Appendix Table 6**), which suggested that SGLT2 inhibition is likely to show an effect on reducing risk rather than influencing the diagnostic work up for prostate cancer. As a positive control, we confirmed the well-established effect of SGLT2 inhibition on reducing the risk of T2DM (OR=0.66, 95%CI=0.49 to 0.88, P=0.005; **Appendix Table 6**).

The validation MR analysis using the two instruments selected by the stringent approach and using SGLT2 instruments derived from the MAGIC consortium validated the effect of SGLT2 inhibition on total, advanced and advanced vs localised prostate cancer (**Figure 3** and **Appendix Table 7**).

#### Tests of MR assumptions

The exchangeability MR assumption was tested using genetic colocalization between SGLT2 inhibition and prostate cancer, where we observed evidence of colocalization of the two traits in the *SLC5A2* region (colocalization probability=72%; **Appendix Table 8**).

The exclusion restriction MR assumption was examined in several analyses. The PheWAS of the primary SGLT2 instruments showed that these genetic variants were associated with blood cell traits (e.g. red blood cell counts), body weight traits (e.g. waist circumference), diastolic blood pressure and low-density lipoprotein cholesterol (**Appendix Table 9**). Multivariable MR adjusting for these traits respectively (**Appendix Table 10A**) suggested that the effect of SGLT2 inhibition on prostate cancer was independent of these traits (**Appendix Table 10B**). In addition, the SGLT2 instruments showed associations with the expression of 17 genes excluding *SLC5A2*, with two genes being targets for existing drugs for coagulation and hemoglobinuria treatment. The 17 genes were not associated with glycemic traits or to have an interaction with any anti-diabetic or anti-cancer drugs^50^ (**Appendix Table 11**). The differential expression analysis using data from the TCGA Program showed little evidence of differential expression levels in prostate tumour tissue versus normal tissue for 15 of the 17 genes (except *SLC5A2*) (**Appendix Table 12**), implying that pleiotropy was less likely occur via these genes.

The MR sensitivity analyses did not provide strong evidence of heterogeneity or pleiotropy for the effect of SGLT2 inhibition on prostate cancer, but the statistical power to clearly demonstrate this was low (**Appendix Table 6** and **7**).

### Validation of association of usage of SGLT2 inhibitors with prostate cancer risk using electronic healthcare data

We identified 26,988 new users of SGLT2 inhibitors and 54,134 new users of DPP4 inhibitors who fulfilled the eligibility criteria out of 130,817 males from SLHD (**Figure 2A**). After a 1:1 propensity-score matching, we identified a cohort of 48,310 patients (24,155 in each group) with well-balanced baseline characteristics (standardized mean differences less than 1.5%) between the two treatment groups (**Appendix Table 5A**). Cox proportional hazards model showed that SGLT2 inhibitors use (compared with DPP4 inhibitors use) was associated with a 23% reduction in risk of prostate cancer (SGLT2 inhibitors use=467.4 versus DPP4 inhibitors use=492.75 per 100,000 person-years; HR=0.77, 95%CI=0.61 to 0.99, P=0.03) during a median follow-up of 1.33 years (**Figure 2B**). Sensitivity analyses lagging the outcome period between one to six months showed similar protective effects, albeit less precisely estimated (**Appendix Table 13**).

### Biological validation: association of *SLC5A2* expression on prostate cancer

Differential expression analysis showed that the expression levels of *SLC5A2* was 2.02 folds higher in prostate cancer tissue than in normal prostate tissue (P=0.006; **Figure 4**). Two additional genes, *TGFB1I1* and *ITGAD,* also showed differential expression in normal versus tumour tissues (*TGFB1I1*: log2FoldChange=-1.65, P=1.95×10^-7^; *ITGAD*: log2FoldChange=1.47, P=0.003; **Appendix Table 12**). Although these two genes could be considered as potential anti-prostate cancer drug target, they were not reported to be associated with any diabetic or glycemic traits or considered as anti-diabetic drugs. Therefore, they were not likely to be pleiotropic exposures that linking SGLT2 instruments with prostate cancer.

**Figure 4.**
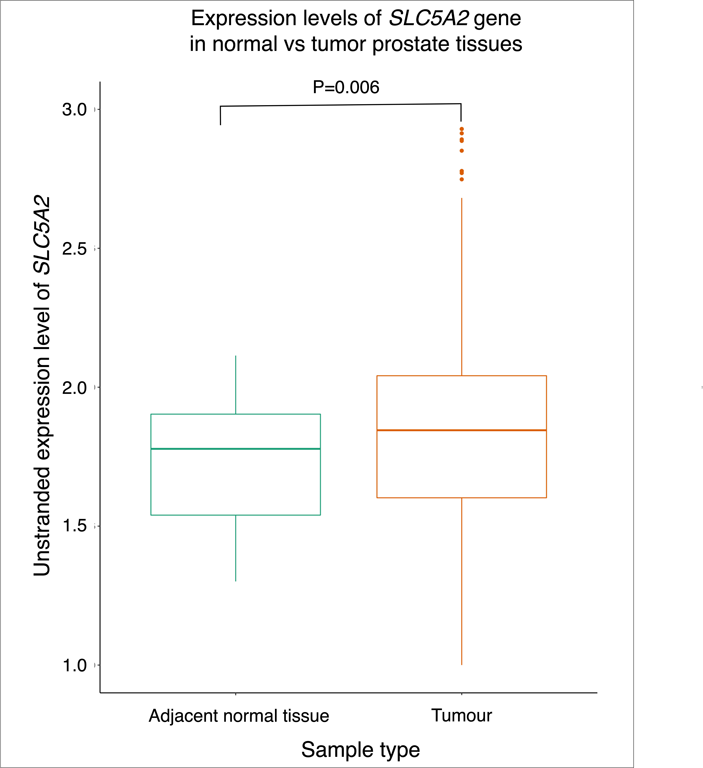
Differential expression analysis results of *SLC5A2* expression in prostate tumor vs adjacent normal tissue.

### Validating the influence of glucose: MR and observational association of HbA_1c_ with prostate cancer

We estimated the association of HbA_1c_ with prostate cancer risk using MR and observational analyses, which aimed to investigate whether the effect of SGLT2 inhibition on prostate cancer is partly via lowering HbA_1c_ levels. Little evidence was observed to support the effect of genetically proxied HbA_1c_ on change prostate cancer risk (OR=0.98, 95%CI=0.92 to 1.05, P=0.63; **Table 3**). Sensitivity MR analyses in which we removed variants within the *SLC5A2* region showed similar effects to those seen in our analyses of HbA_1c_ on prostate cancer (**Appendix Table 14**). Observational analysis in the REACTION study also provided little evidence to support the effect of baseline HbA_1c_ levels on incident prostate cancer after 10 years of follow-up (HR=0.93, 95%CI=0.80 to 1.10, P=0.40), the findings barely change after excluding individuals using anti-diabetic drugs (**Table 3**).

**Table 3.**
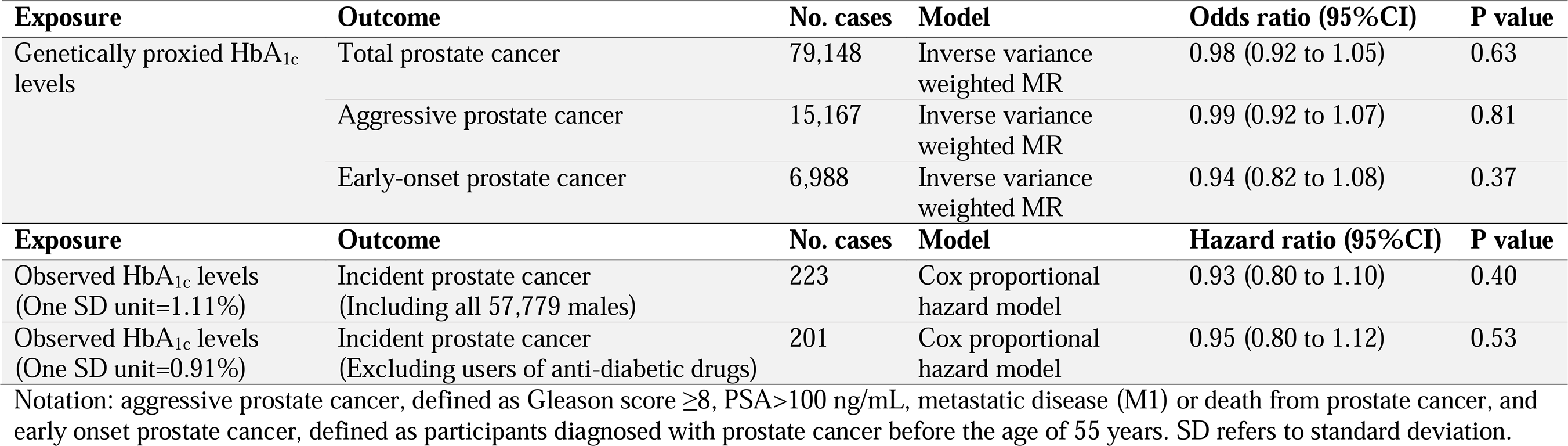
Effect estimates of genetic proxied HbA_1c_ levels on total, aggressive and early onset prostate cancer among men in general population using data from PRACTICAL consortium and effects of observed HbA_1c_ levels on incident prostate cancer among men in general population using data from the REACTION study.

## Discussion

In this study, we triangulated human genetics, electronic healthcare and biological evidence to answer the same causal question: the effect of SGLT2 inhibition on prostate cancer. In the discovery, we observed that genetically proxied lifelong SGLT2 inhibition reduced total-, advanced- and early-onset prostate cancer in the general male population by 44%, 48% and 73%, respectively. Validation using various selection processes and datasets confirmed the protective effect of SGLT2 inhibition on risk of prostate cancer and its subtypes, rather than an effect on PSA biasing the diagnosis of prostate cancer. In the validation using electronic healthcare data, we showed that SGLT2 inhibitor use reduced risk of prostate cancer by 23% in men with T2DM. In the biological validation, we showed that expression levels of SGLT2 were 2.02 times higher in prostate tumour tissue than that in normal prostate tissue. In the analyses validating the influence of glucose, we found little genetic and observational evidence to support an association of HbA_1c_ on prostate cancer, which implies a possible non-glucose mechanism of SGLT2 inhibition on prostate cancer prevention (see **Appendix Figure 3**). Correctively, we provide three strands of evidence to prioritise SGLT2 inhibition as a target for prostate cancer prevention and adjunctive treatment.

According to the U.S. Centers for Disease Control and Prevention (CDC), adults aged between 45 and 64 receive the greatest number of new diagnoses of diabetes, which was also the age group that men are likely to receive diagnoses of prostate cancer. However, there was little evidence to support setting up of clinical guidelines concerning modification of SGLT2 inhibitor treatment among diabetic patients with co-existing prostate cancer till now. A few epidemiology studies supported the protective role of SGLT2 inhibitors on prostate cancer risk^13,51^. A recent systematic review of RCTs provided weak evidence of an effect of SGLT2 inhibitors on cancers^52^. Only one phase I trial was registered in clinicaltrial.gov (NCT04887935), which aims to investigate the safety of dapagliflozin, one type of SGLT2 inhibitors, for men considered at high risk of prostate cancer. In the present study, we observed robust human genetics and electronic healthcare evidence to support the effect of SGLT2 inhibition on reducing risk of prostate cancer, both in the general male population and in males with diabetes. Our results further support that SGLT2 inhibition may have better efficacy on prevention of early-onset prostate cancer than on total- and advanced prostate cancer. Our evidence supports the prioritization of future clinical trials of SGLT2 inhibitors in diabetic men at high risk of prostate cancer, which may have the potential to influence clinical guidelines/standards for diabetes.

It has been hypothesized that the primary mechanism of a beneficial effect of SGLT2 inhibitors on cancer is through inhibiting glycolysis in tumor cells, thus reducing tumor cell proliferation and tumorigenesis^53^. Another study showed that Canagliflozin, one type of SGLT2 inhibitor, inhibits mitochondrial complex-I and cellular proliferation in prostate cancer cells^54^. However, the lack of MR and observational evidence of a role for HbA_1c_^29^ suggests that HbA_1c_ may not be driving the observed association of SGLT2 inhibition with prostate cancer. Recent *in-vitro* and *in-vivo* studies have shown that SGLT2 antibodies are expressed in prostate adenocarcinomas but are rarely expressed in normal prostate cells surrounding cancer cells^55^. But these studies were conducted in a small number of prostate samples, raising the question of generalizability of the findings. In our study, we harmonized data from over 600 prostate tumour tissues and over 50 normal prostate tissues. Our analysis supports an association of *SLC5A2* expression and prostate cancer in males with prostate cancer. Correctively, our genetic and biological evidence imply that SGLT2 inhibition may have a direct effect on prostate cancer prevention and treatment, which could be independent to its glucose control effect. Some well-designed clinical trials have also provided evidence to support that SGLT2 inhibitors have good tolerance and safety profiles to be used in individuals without diabetes^56,57^. Further functional and clinical studies are warranted to better understand the anti-cancer mechanism of SGLT2 inhibitors and test their anti-prostate cancer efficacy in individuals without diabetes.

### Strengths and limitations

Our study has several strengths. First, we estimated the effects of SGLT2 inhibition on prostate cancer prevention and treatment using genetic, epidemiological and biological approaches, which have different assumptions, key source of biases (e.g. pleiotropy for MR and confounders for observational analysis)^15^ and different subgroup of population (i.e. the general male population, males with diabetes and males with prostate cancer). Triangulation of evidence suggests that SGLT2 inhibition is likely to have a protective effect on prostate cancer in all subpopulation groups, which strengthens confidence in this finding. Second, the instruments for SGLT2 were selected using two widely applied pipelines^18,29^. The reliability of these instruments has been tested thoroughly in this study. Third, we paid special attention to the potential influence of our genetic variant-exposure estimates on our MR results and only used male-specific instruments in this study. Fourth, the results from colocalization analysis, PheWAS, multivariable MR, and other sensitivity MR analyses suggested that the effect of SGLT2 inhibition on prostate cancer is unlikely to violate the exchangeability and the exclusion restriction assumptions of MR. More interestingly, we extended the scope of differential gene expression analysis to distinguish pleiotropy from causality, which the strategy can be widely applied to other drug target genes and complex diseases.

This study has several limitations. First, our MR estimates of the effect of SGLT2 inhibition were scaled to represent the on-target reductions in HbA_1c_ levels rather than the direct effect of SGLT2 inhibitors. This assumes that SGLT2 inhibition has a proportional impact on lowering of HbA_1c_. Second, caution is needed to interpret the causal effect estimate from this study. This is because the MR estimate reflects the long-term modulation of drug targets on disease risk, which may suggest different levels of risk reductions per unit change in drug target compared with those observed from clinical trials/observational studies over a relatively short duration, which would explain the attenuated effect estimate of our observational analysis. Furthermore, the estimated effect of SGLT2 inhibition on prostate cancer could at least in part be influenced by different ancestries, disease status and survival bias given the relatively late age-at-onset of prostate cancer. Fourth, the MR analyses presented assumes no gene-environment interaction of the association of genetic proxies for drug targets and prostate cancer. Fifth, SGLT2 inhibitors have been marketed in China since March 2017, the median follow-up time for the observational analysis was therefore only 1.33 years. Therefore, we consider this result as a validation for evidence triangulation rather than a stand-alone finding.

## Conclusions

Genetic, epidemiological, and biological evidence with different assumptions and used different subpopulations support the role of SGLT2 inhibition on reducing prostate cancer risk. Further clinical trials should be prioritised to establish whether there is a similar effect with the long-term prescription of SGLT2 inhibitors, at what age chemoprevention/treatment would need to commence, whether high-risk men should be targeted and the potential harms.

## Data Availability

The data, analytic methods, and study materials will be made available to other researchers for purposes of reproducing the results. In more details, the genetic association data of the selected risk factors are available in Appendix Tables. The summary level GWAS statistics for the primary and secondary outcomes are available from the MRC IEU OpenGWAS database https://gwas.mrcieu.ac.uk/ UK Biobank received ethical approval from the Research Ethics Committee (REC reference for UK Biobank is 11/NW/0382). The analytical script of the MR analysis that had been used in this study is available via the GitHub repository of the TwoSampleMR R package https://github.com/MRCIEU/TwoSampleMR/

## Acknowledgements

J.Z. is supported by the Academy of Medical Sciences (AMS) Springboard Award, the Wellcome Trust, the Government Department of Business, Energy and Industrial Strategy (BEIS), the British Heart Foundation and Diabetes UK (SBF006\1117). J.Z. is funded by the Vice-Chancellor Fellowship from the University of Bristol. J.L.L., M.X., G.N., and Y.B. are supported by the National Natural Science Foundation of China (82088102, 81970691, 81970728, 81930021 and 81941017) and Shanghai Clinical Research Center for Metabolic Disease (19MC1910100). J.L.L., M.X., Y.X., T.W., M.L., Z.Z., S.W., H.L., G.N., W.W., and Y.B. are members of the Innovative Research Team of High-level Local Universities in Shanghai. G.N. and Y.B. are supported by the Shanghai Shenkang Hospital Development Center (SHDC12019101, SHDC2020CR1001A, SHDC2020CR3064B). V.W., G.D.S. and T.R.G. are supported by the UK Medical Research Council Integrative Epidemiology Unit (MC_UU_00032/03). R.M.M, S.H. and P.D. received support from a Cancer Research UK (C18281/A29019) programme grant (the Integrative Cancer Epidemiology Grant). R.M.M., T.R.G. & G.D.S. are also supported by the NIHR Bristol Biomedical Research Centre which is funded by the NIHR and is a partnership between University Hospitals Bristol and Weston NHS Foundation Trust and the University of Bristol. Department of Health and Social Care disclaimer: The views expressed are those of the author(s) and not necessarily those of the NHS, the NIHR or the Department of Health and Social Care. JY is supported by a Cancer Research UK Population Research Postdoctoral Fellowship (C68933/A28534). R.M.M. is a National Institute for Health Research Senior Investigator (NIHR202411). GDS reports Scientific Advisory Board Membership for Relation Therapeutics and Insitro.

We thank the individual patients who provided the sample that made data available; without them the study would not have been possible. We thank for PRACTICAL and GAME-ON/ELLIPSE consortia provided key genetic data to support this study.

## Author contributions

J.Z., G.N., R.M., W.W., and Y.B. designed the study, wrote the research plan, and interpreted the results. J.Z. undertook the main, replication and sensitivity MR analyses with feedback from Y.Q., O.D., J.Y., and J.R. B.C. and J.Q. collected data from the Shanghai Link Healthcare Database and conducted the survival and linear regression analyses, with C.S.L.C., S.L.A.Y., S.L., and J.Y. provided critical suggestions. H.L.Z. conducted the computational biology analyses using TCGA, with supports from Y.Y.Q. J.Z. and J.L. wrote the first draft of the manuscript with critical comments and revision from M.X., Y.X., T.W., M.L., Z.Z., R.Z., S.W., H.L., C.Y.H., C.S.L.C., S.L.A.Y., S.L., O.D., P.D., S.H., Y.L., J.R., J.Y., P.H., J.Y., S.L., T.R.G., G.D.S., R.M., W.W., Y.B., and G.N. J.Z. is the guarantor. The corresponding author attests that all listed authors meet authorship criteria and that no others meeting the criteria have been omitted.

## Data and materials sharing

The data, analytic methods, and study materials will be made available to other researchers for purposes of reproducing the results. In more details, the genetic association data of the selected risk factors are available in Appendix Tables. The summary level GWAS statistics for the primary and secondary outcomes are available from the MRC IEU OpenGWAS database (https://gwas.mrcieu.ac.uk/). UK Biobank received ethical approval from the Research Ethics Committee (REC reference for UK Biobank is 11/NW/0382). The analytical script of the MR analysis that had been used in this study is available via the GitHub repository of the TwoSampleMR R package (https://github.com/MRCIEU/TwoSampleMR/).

## Ethical approval

UK Biobank has received ethical approval from the UK National Health Service’s National Research Ethics Service (ref 11/ NW/0382). All other studies contributing data to this analysis had the relevant institutional review board approval from each country and all participants provided informed consent.

## Transparency

The lead author (J.Z.) affirms that this manuscript is an honest, accurate, and transparent account of the study being reported; that no important aspects of the study have been omitted; and that any discrepancies from the study as planned (and, if relevant, registered) have been explained.

