## Supplementary Figures and notes for "Mendelian randomization revealing the protective effect of sodium-glucose cotransporter 2 inhibition on prostate cancer with verified evidence from electronic healthcare and biological data"

### Supplementary appendix

**Appendix Figure 1. Three sets of instruments been used in the Mendelian randomization analysis.**

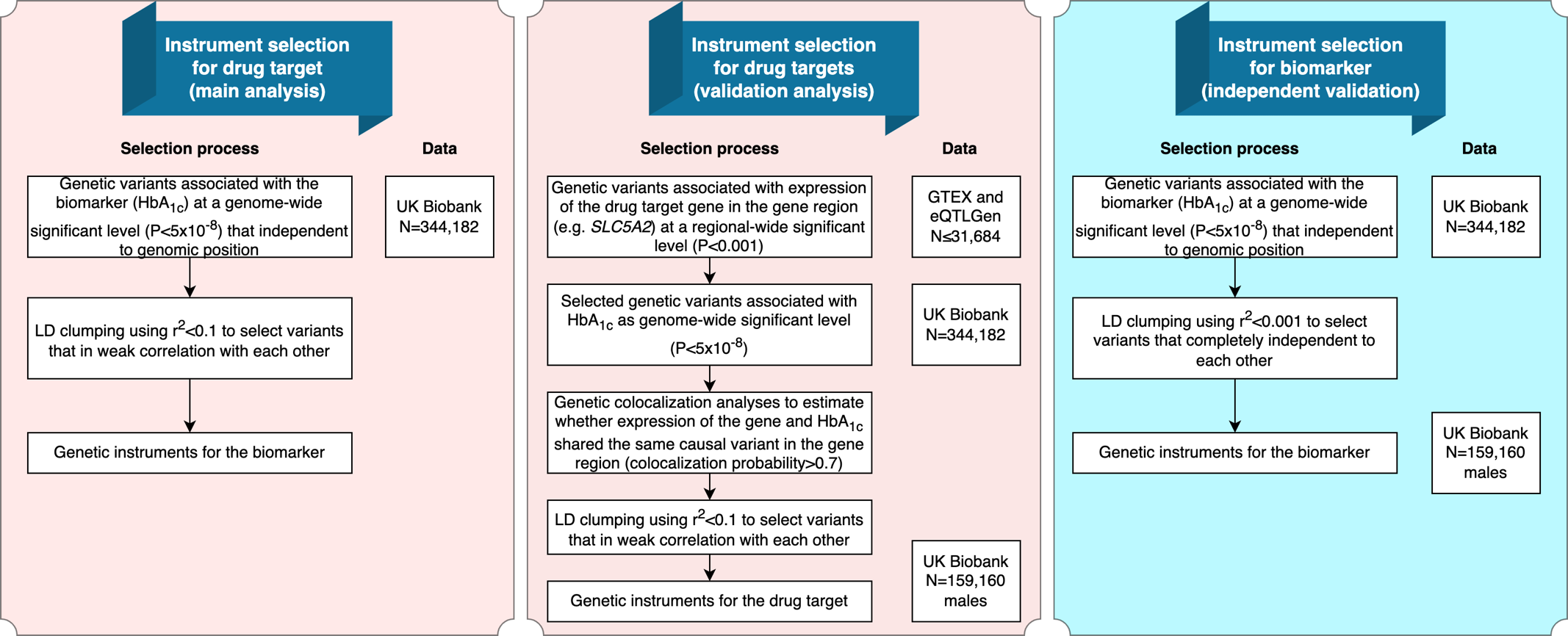

**Appendix Figure 2. Scatter plot and forest plot for the genetically proxied effect of SGLT2 inhibition on total prostate cancer.**

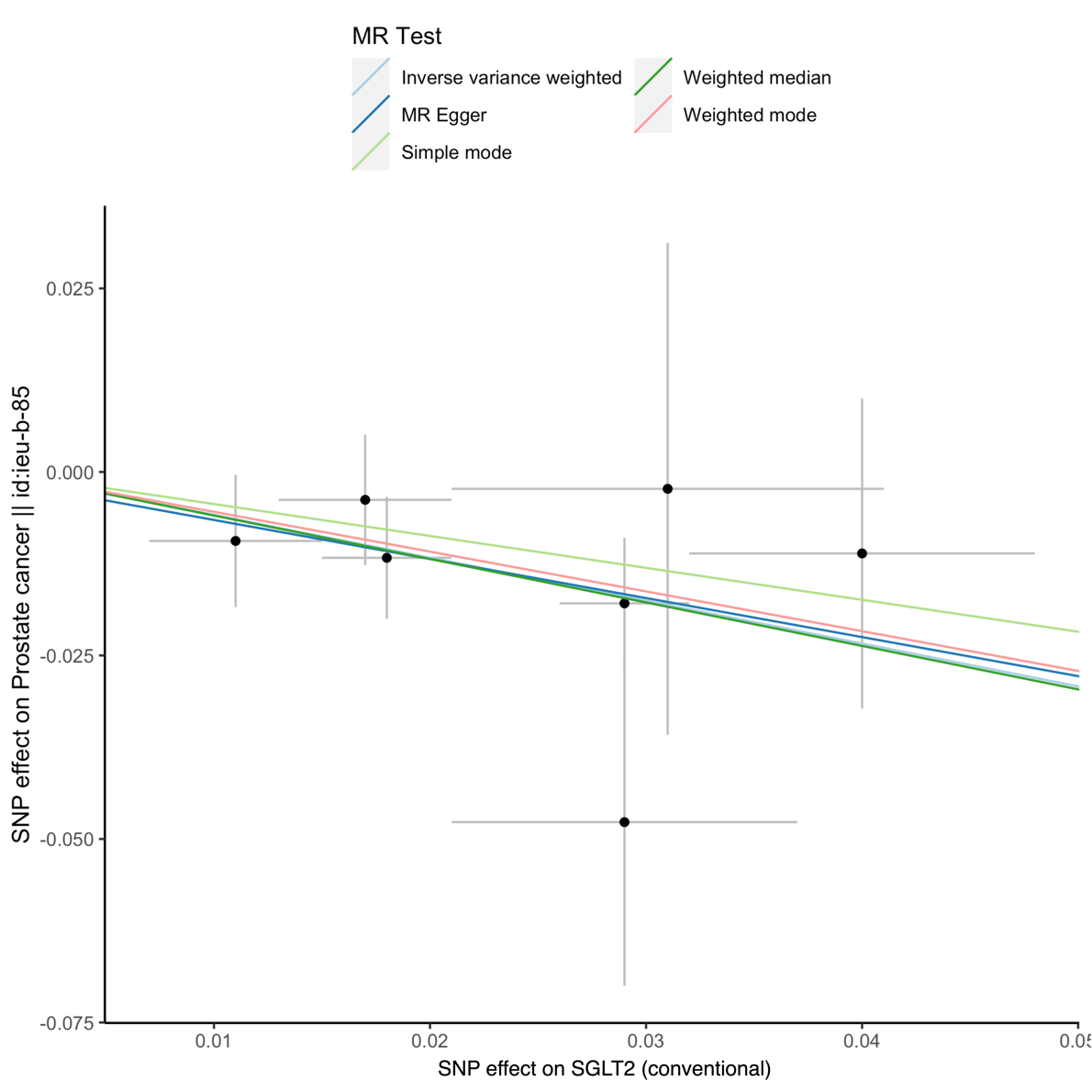

**Appendix Figure 3. Summary of key findings of the current study.**

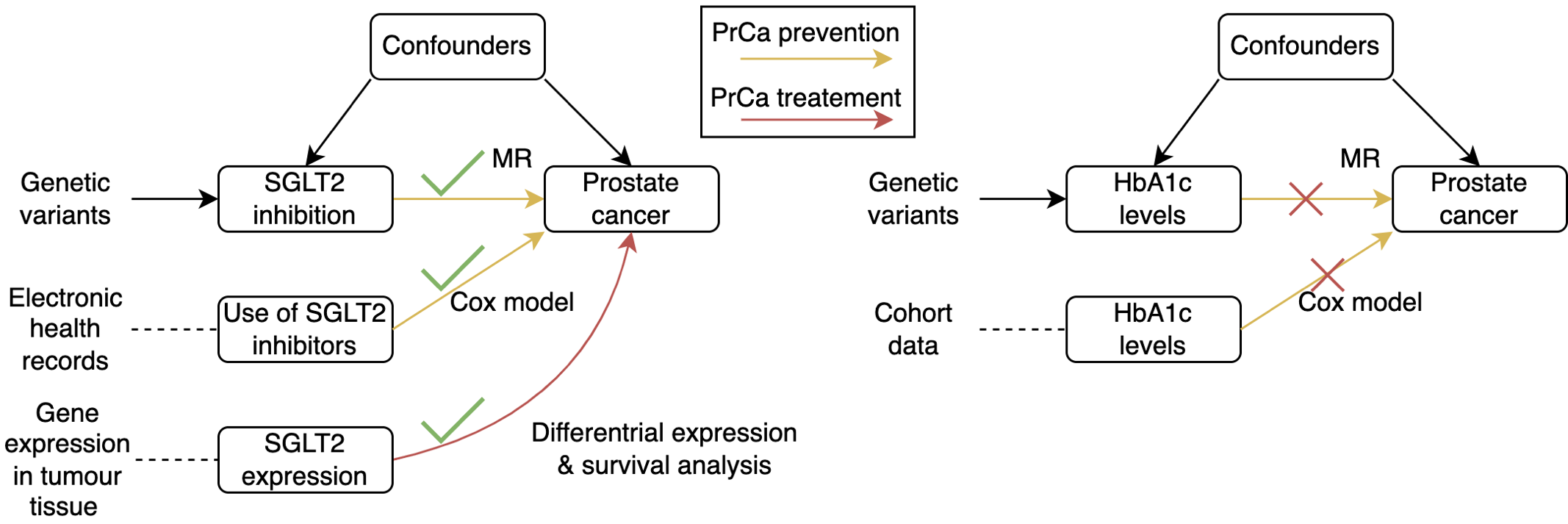

### Appendix Note 1. Details of instrument selection

**Identification of drug target of SGLT2 and exposure data**

This study investigated drug target for SGLT2 inhibitors. The drug targeted gene of for SGLT2, *SLC5A2* was well defined in the literature^1^.

Three sets of genetic instruments were used to proxy effect of SGLT2 inhibition (**Supplementary** **Figure 1**). For main drug target MR, summary data were obtained from a GWAS of HbA_1c_ levels in the UK Biobank (N=159,160 males), in which genetic variants associated with HbA1c in the SGLT2 region were selected as instruments. For the validation MR, a set of genetic variants associated with both HbA1c and expression levels of SGLT2 (data from the GTEX and eQTLGen consortia [N≤31,684]^2,3^).

For independent validation MR analyses, the GWAS of HbA_1c_ levels from the MAGIC consortium^4^ were used. The primary MAGIC GWAS was a trans-ancestry meta-analysis, for which we consider population structure may be a confounder to bias the MR estimates. We therefore used the European-only GWAS results from 146,806 European individuals. In addition, since the genetic effects of the MAGIC HbA_1c_ GWAS was scaled to percentage unit in the original study. We conducted a beta transformation for the genetic effects of HbA_1c_. After transformation, the unit of HbA_1c_ GWAS was changed to standard deviation (SD) decreasing unit. By applying this transformation, the MR effect estimates were comparable between UK Biobank and MAGIC. In addition, For the MAGIC GWAS, individuals with type 1 or type 2 diabetes, with usage of diabetes-relevant medications or has a fasting glucose 7 mmol l^−1^, 2-hour glucose ≥ 11.1 mmol l^−1^ or HbA_1c_ ≥ 6.5% were excluded from the analysis.

**Instrument selection**

As demonstrated in **Supplementary** **Figure 1**, we applied three instrument selection approaches to select genetic instruments for SGLT2 inhibition from two independent datasets.

The first approach selected SGLT2 instruments from a classic drug target instrument selection process (primary instruments). The genetic variants associated with HbA_1c_ with a genome-wide association threshold of P<5×10^-8^ in the *SLC5A2* gene region (target gene for SGLT2 inhibition) were selected as candidate instruments. After selection, seven variants that proxying SGLT2 inhibition were selected as set 2 instruments for SGLT2 inhibition (**Supplementary** **Table 1**).

The second approach selected instruments for the main drug target MR analyses (stringent instruments). Genetic variants associated with expression levels of drug target genes in a regional-wide significance threshold (P<0.001) and HbA_1c_ in a genome-wide significance level (P<5×10^-8^) in a genomic region near the drug target gene (±1Mb window) were selected as candidate instruments. We systematically scanned genetic variants associated with the expression levels of *SLC5A2* using data from seven recent GWAS studies of genes level in 49 human tissues and proteins in plasma^5,6,7,8,9,10,11^. This is because targets for SGLT2 inhibition may influence glycemic traits via biological mechanisms in different tissues. A set of genetic colocalization methods^12,13^ were then used to select genetic variants with shared causal variants of expression level of the drug target gene and HbA_1c_ in the gene coding region. This step mapped 44 genetic variants for SGLT2 (**Supplementary Table 2**). We further applied linkage disequilibrium (LD) clumping to select those with the lowest P value that had a LD (which refers to pairwise squared correlation [r^2^]) less than 0.1 as this indicates weak correlation among the selected genetic variants. European population specific LD among variants were estimated from the 1000 Genomes Project (phase 3) implemented in the two-sample MR package^15,14^. After filtering, two variants were selected as instruments for SGLT2 inhibition (**Supplementary** **Table 1**).

The third approach selected instruments of SGLT2 inhibition from an independent dataset from MAGIC consortium. The genetic variants passed regional-wide association threshold of P<1×10^-5^ in the *SLC5A2* region were selected as candidate instruments. LD clumping with a threshold of 0.01 was further applied to select complete independent genetic variants as genetic instruments. After selection, one genetic variant that proxying SGLT2 inhibition were selected as instrument (**Supplementary** **Table 3**).

### Appendix Note 2. The observational analysis in the Shanghai Link Healthcare Database and the REACTION study

**The survival analysis of use of SGLT2 inhibitors on prostate cancer/high risk of prostate cancer in the Shanghai Link Healthcare Database**

The survival analysis was conducted using data from the Shanghai Link Healthcare Database (SLHD)^15^, a representative clinical database covering > 99% of Shanghai residents. The SLHD is developed and operated by the Shanghai Hospital Development Center (SHDC), which is an administrative department of the Shanghai Municipal Government. The SHDC is responsible for the surveillance of 35 tertiary hospitals in Shanghai. In China, government-run hospitals are classified as primary (grade I), secondary (grade II), or tertiary (grade III) hospitals according to their abilities in medical care, medical education, and medical research, with tertiary hospitals being the best. According to administrative regulations, all 35 tertiary hospitals are required to upload general medical practice data (i.e., outpatient visits, emergency department visits, and hospital admissions) to the SLHD. Any personally identifiable information is scrambled to protect privacy. The SLHD has released data for academic research since 2013, which requires review and approval to access.

**Figure 2A** illustrates the selection process of the study population. First, all males aged between 40-99 years newly treated with SGLT2 inhibitors or DPP4 inhibitors from March 1, 2017 to December 31, 2021 were identified. Cohort entry was defined as the date of the first prescription. Exclusion criteria were defined as follows: patients without any medical record before cohort entry; patients who had been treated with both SGLT2 inhibitors and DPP4 inhibitors; patients with a history of prostate cancer; patients with total prostate specific antigen (PSA) > 10 ng/mL prior to enrolment; patients with less than 1 day of follow-up. All patients were followed until diagnosis of prostate cancer or death, or December 31, 2021, whichever occurred first.

The following covariates that may affect prostate cancer risk and/or total PSA levels were adjusted in the cox model: demographic data included age; comorbidities included benign prostatic hyperplasia, hypertension, dyslipidemia, diabetic complications, ischemic heart disease, peripheral vascular disease, heart failure, cerebrovascular disease, chronic lung disease, moderate or severe kidney disease, moderate or severe liver disease, and other cancers; antidiabetic drugs included metformin, insulin, glucagon-like peptide-1 receptor agonist, sulfonylurea, glinide, α-glucosidase inhibitor, and thiazolidinedione; other medications included angiotensin converting enzyme inhibitor, angiotensin receptor blocker, calcium channel blocker, α/β-blockers, diuretic, statin, fibrate, aspirin, other antiplatelet drugs, non-steroidal anti-inflammatory drug, and 5α-reductase inhibitor. All comorbidities and medications records were assessed by relevant medical records prior to cohort entry. In addition to the original cohort, we also established a 1:1 propensity score matched cohort of SGLT2 inhibitors users and DPP4 inhibitors users (caliper: 0.20 standard deviation of the logit of the estimated propensity score). Standardized mean differences (SMDs) were calculated for all covariates between SGLT2 inhibitors users and DPP4 inhibitors users, with values less than 10% likely to indicate relative balance.

For the survival analysis, baseline characteristics of SGLT2 inhibitors users and DPP4 inhibitors users are presented as medians with interquartile ranges (IQRs) for continuous variables and frequencies with percentages for categorical variables. The crude incidence rate of prostate cancer-by-proxy was calculated by dividing the number of cases by the number of person-years. Cox proportional hazards models were used to estimate hazard ratios (HRs) and 95% confidence intervals (CIs) of incident prostate cancer-by-proxy, comparing SGLT2 inhibitors use with DPP4 inhibitors use. Sensitivity analyses were performed by setting different lag periods: 1-month, 2-month, 3-month, and 6-month lag period. Statistical analyses were performed using R language software (version 4.1.2).

**The prospective association of HbA_1c_ with incident prostate cancer in the Risk Evaluation of Cancers in Chinese diabetic individuals: a longitudinal (REACTION) Study**

We estimated the association between HbA_1c_ and incident prostate cancer during a median of 10.1 years of follow-up in the Risk Evaluation of Cancers in Chinese diabetic individuals: a longitudinal (REACTION) study^16,17,18,19,20^. The REACTION study was a multicenter, population-based, prospective cohort study aiming to demonstrate whether abnormal glucose metabolism (diabetes and prediabetes) was associated with increased risk for cancer in the Chinese population and to identify factors that modify the risk of cancer among individuals with abnormal glucose metabolism. Between 2011 and 2012, a total of 259657 individuals aged 40 years and older were recruited from 25 communities of various regions of China. Eligible men and women aged ≥40 years were identified from local resident registration systems. Trained community health workers visited eligible individuals’ homes and invited them to participate in the study. Due to limited funds, a total of 193846 participants of 20 communities from REACTION were followed up during 2021-2022. After excluding participants with prostate cancer at baseline, we included 57,779 men aged 40 years or older in the current analysis.

The study was approved by the Medical Ethics Committee of Ruijin Hospital, Shanghai Jiao-Tong University. All study participants provided written informed consent.

As described previously^16,17,18,19,20^ HbA_1c_ was determined by using high-performance liquid chromatography (VARIANT II System; Bio-Rad Laboratories) in the central laboratory located at Ruijin Hospital, Shanghai, China, which is certificated by the U.S. National Glycohemoglobin Standardization Program and passed the Laboratory Accreditation Program of the College of American Pathologists. Information on prostate cancer were collected from local death and disease registries of the National Disease Surveillance Point System and National Health Insurance System with use of the ICD 10 code “C61” in the study. Cox proportional hazards model was applied to estimate the hazard ratio of HbA_1c_ on incident prostate cancer in the overall population (N=57,779). A sensitivity analysis was performed in participants without receiving glucose-lowering therapy at baseline (N=53,037). Age, body mass index, tobacco consumption, alcohol consumption, physical activity, and diet score were included as covariates in the model.

### Appendix Note 3. Description of MR sensitivity analyses

The MR assumptions were tested using the following sensitivity methods.

MR exploits both Mendel’s Law of Heredity^21^. The Law of Independent Assortment refers to the fact that alleles of genes in different parts of the genome are inherited independently. Compliance with this Law was evaluated using a generalized inverse variance weighted (gIVW) model^22^, which takes into account the weak LD (r^2^=0.089) between the SGLT2 instruments.

The MR assumption of relevance was tested by generating estimates of the proportion of variance in each drug target explained by the instrument (R^2^) and F statistics. An F statistic of at least 10 is indicative of evidence against weak instrument bias (a reduction in statistical power to reject the null hypothesis when an instrument explains only a small proportion of variance in an exposure)^23^.

The MR assumption of exchangeability was tested by performing a genetic colocalization analysis between the drug target and prostate cancer^12,13^. This can be used to assess whether false-positive drug target-disease associations were created due to confounding by LD between nearby genetic variants (genetic confounding). A posterior probability of colocalization over 70% between a drug target and prostate cancer was used as evidence of colocalization.

The MR assumption involving the exclusion restriction was tested using a whole set of sensitivity methods. First, the presence of an association between an instrument for SGLT2 inhibition and an off-target phenotype could provide evidence of horizontal pleiotropy (which means a genetic variant influences a phenotype through biological pathways that are independent of the exposure under investigation), which is a violation of the exclusion restriction criterion. A phenome-wide association study (PheWAS) of the genetic instruments for SGLT2 inhibition was performed among a comprehensive list of 22,479 human phenotypes included in the IEU OpenGWAS database^41,24^. If there was evidence of effect of genetic instruments for SGLT2 inhibition with unintended phenotypes at a genetic association threshold of 5×10^-8^, multivariable analyses were performed to examine associations between the genetic instruments for SGLT2 inhibition and prostate cancer outcomes, adjusted for genetically proxied phenotype^25^.

Second, if there was evidence of genetic effect of the SGLT2 instruments on expression levels of other genes, where the expression levels of these genes were associated with prostate cancer, then this will violate the exclusion restriction assumption of MR. We therefore conducted a transcriptome wide variant lookup to identify all genes that are associated with the SGLT2 instruments with P<1×10^-4^ (**Supplementary Table 11**). Differential expression analysis was then applied for expression levels of these genes in prostate tumour tissue versus normal prostate tissue. If expression level did not different between the two tissues, we will be more confident that these genes are not likely to be pleiotropic exposures that linking SGLT2 instruments with prostate cancer risk.

Third, the violations of the exclusion restriction assumption were further tested by examining associations of the genetic instruments with four previously reported causal prostate cancer risk factors (accelerometer-based physical activity measurement, serum iron, body mass index and monounsaturated fatty acids)^26^. A marginal MR threshold (P<0.05) was used as evidence of a potential pleitropy effect of the genetic instruments for SGLT2 inhibition on prostate cancer via a prostate cancer risk factor.

Fourth, for genetic instruments for SGLT2 inhibition with two or more SNPs, evidence of horizontal pleiotropy was examined via the following sensitivity analyses: heterogeneity test across instruments using Cochran’s Q and Rücker’s Q^27,28^, weighted median^29^ and mode-based estimate approaches^30^. Weighted median MR and mode estimator approaches^29,30^ are two additional sensitivity analyses, which provide consistent causal estimates of the exposure on the outcome even when up to 50% (or up to 100% for the mode estimator approach) of the information contributing to the analysis comes from genetic variants that exhibit pleiotropy (or even the majority of information in the case of the mode-based MR).

If all MR sensitivity methods provide similar causal estimates of genetic proxied SGLT2 inhibition on prostate cancer, we are more confident that the causal estimates were robust to various MR assumptions.

### Appendix Note 4. Description of differential expression analysis and survival analysis

**Differential expression analysis**

The differential expression analysis was conducted using data from the TCGA-PRAD project. The expression data (raw counts) and tissue type information (tumour or normal solid tissue) of 18 selected genes were downloaded from GDC data portal (refers to tissue samples obtained from prostate cancer patients). After selection, 691 prostate samples for 639 individuals were selected from three databases, TCGA, DTB and PC project, in which 639 were prostate tumour samples and 52 were normal solid tissues.

R package “Deseq2” ^31^ was used to detect differentially expressed genes between tumor and normal tissues. We applied the default normalization and differential expression analysis method using Deseq2^31^, which estimated size factors and dispersions, and then conductd Negative Binomial GLM fitting and calculated Wald statistics. Genes with differential expression log2(FC) > 1 and P_adjusted_<0.05 were defined as showing differential expression between tumour and normal prostate tissues.
